# Research on the Structural Characteristics and Evolution of China’s Medical and Health Innovation Cooperation Network Based on the Chinese Medical Science and Technology Awards

**DOI:** 10.1101/2024.11.21.24317696

**Authors:** Li Xiaoqian, Zhang Fan, Zhang Junliang, Wang Zhengfang, Yu Yue, Chen Huizheng

## Abstract

Cooperative innovation is an important means to promote the development of medical and health undertakings. Based on the award data of the Chinese Medical Science and Technology Award from 2003 to 2022, this paper constructs an innovation cooperation network and uses social network analysis to explore the structure and evolution characteristics of China’s medical and health innovation cooperation network. The research shows that China’s medical and health innovation cooperation continues to increase, but the development of regional cooperation is not balanced; the scale of the network is expanding, and the small world effect is obvious, but the network density is relatively low, and there is still a large space for development of institutional innovation cooperation; Hospitals and universities play an important role in the network; The medical and health innovation cooperation network has the characteristics of community. Finally, policy suggestions are put forward from the aspects of strengthening regional balanced development, giving play to the radiation role of core institutions in the innovation cooperative network, and promoting diversified innovation cooperation, to form a balanced development innovation cooperation system, a robust innovation cooperation network ecology, and a stable innovation cooperation network structure.

## Introduction

In the current knowledge economy era, the high complexity of innovation activities makes it difficult for a single organization to master all the resources and complete all the innovation activities independently[1]. Therefore, innovation cooperation has gradually become the most effective and dynamic driving force to promote scientific and technological innovation[2–3]. As an important part of the national innovation system, innovation cooperation networks can provide practical carriers, environmental support and mechanism guarantees for the development of innovation-driven development [4], and related research has gradually become a hot issue in domestic and international research [5].

### Related work

Countries all over the world attach great importance to scientific and technological innovation in the field of healthcare, and the report of the 20th CPC National Congress puts forward: "Adhere to the scientific and technological frontiers, the main battlefield of the economy, major national needs, and the life and health of the people, and accelerate the realization of high-level self-reliance and self-strengthening in science and technology ". The innovation cooperation network in the field of healthcare is of positive significance for promoting the development of healthcare, and scholars at home and abroad are actively carrying out relevant research[6]. Current research has focused on the following three aspects:

### Research on innovative cooperation models

The subjects of innovation cooperation are diverse, and scholars have been actively exploring cooperation modes among subjects from macro and micro subjects. Bender et al. [7] analyzed and explored patterns of institutional collaboration and co-authorship by constructing a network of innovative collaborations for tropical disease research in Germany. Wang et al. [8] selected technology-based small and medium-sized enterprises in the pharmaceutical industry as the research object to analyze different types of patent network cooperation modes. Based on the development history of antimicrobial drugs from 1995 to 2019, Arslan et al. [9] compared and analyzed the two cooperation models of public-private cooperation and private-private cooperation, and found that public-private collaboration is less effective than private-private collaboration.

### Research on the structural characteristics of cooperative networks

Clarifying the structure of innovation cooperation is of positive significance for the in-depth understanding of the current situation and laws of innovation cooperation. Fiori et al. [10] analyzed the technology R&D collaborative networks and the clinical trial R&D collaborative networks, which in turn presented the structural characteristics of different types of innovation cooperation networks in the pharmaceutical industry. By exploring the innovation network structure of the Guangzhou biomedical industry, Zhang et al. [11] found that the innovation cooperation network has a development trend of openness and complexity. Xia et al. [12] found that the emergency drug innovation collaboration network exhibits a sparse structure with many active factions whose members have collaborative advantages

### Research on the influencing factors of innovation cooperation

Exploring the degree of influence of these factors on innovation cooperation is conducive to strengthening innovation cooperation and promoting the healthy development of the medical field. Michelino et al. [13] used patent data to investigate the cooperation between startups and other innovation subjects and found that the higher the level of specialization in the field of knowledge and the higher the level of open innovation, the higher the tendency to innovate and cooperate with scientific research organizations. Lu et al. [14] have demonstrated that the initial time of institutional participation in the innovation cooperation network, the close geographical distance and similar cognitive structure between institutions have a significant impact on the formation of innovation cooperation in the smart healthcare industry.

Innovation cooperation is conducive to promoting scientific and technological innovation in the field of medical and health care, and the innovation cooperation network is an important form of carrier to characterize innovation cooperation. However, the research is mostly gathered in specific fields in medical and health care, and the data sources are mainly patents or journal literature, which makes it difficult to comprehensively analyze the characteristics of the innovation cooperation network in the field of medical and health care and its evolution law from the whole field. Chinese Medical Science and Technology Award rewards units and individuals who have made outstanding contributions to medical science and technology advancement activities, and the form of application results are diversified. Given this, this paper takes the award-winning data of the Chinese Medical Science and Technology Award as the data source and adopts the social network analysis methodology to analyze the network structure and evolution of the medical and health innovation cooperation to comprehensively present China’s medical and healthcare scientific and technological innovation and cooperation network structure, explore its development and evolution characteristics and provide a reference for promoting China’s medical science and technology innovation cooperation.

## Materials and methods

### Data source and data processing

In this paper, we select the award data of the Chinese Medical Science and Technology Award (CMSTA) from 2003 to 2022 to study the innovation cooperation in the field of healthcare in China. And the data comes from the official website of the China Medical Association where a total of 1,782 pieces of award data were obtained.

Before analyzing the data, the data is firstly normalized, and the basic principles are as follows: (1) if the name is changed, its former and current names are counted as the same institution for cooperation; (2) universities and their secondary colleges and research institutes, etc., are counted as the same institution; (3) hospitals, enterprises and the government are counted as independent institutions; and (4) data of awards of single institutions and invalid data, etc., which does not belong to the institutional collaborations, were sifted out, and finally obtain a total of 816 items, accounting for 48.73% of all items.

### Analysis methods and research indicators

Institutional cooperation is when the award-winning innovation projects are jointly accomplished by two or more institutions; intra-regional cooperation is that the institutions of the award-winning innovation projects belong to the same province (municipality directly under the central government, autonomous region, special administrative region) in China; inter-regional cooperation is that the institutions of the award-winning innovation projects belong to different provinces (municipality directly under the central government, autonomous region, special administrative region) in China, or to other countries and regions. The innovation cooperation network is a social network structure formed with innovation institutions as nodes and inter-institutional cooperation as edges.

This paper firstly adopts the statistical analysis method to analyze the cooperation trend and regional cooperation of the award-winning institutions of Chinese Medical Science and Technology Award; secondly, the social network analysis method is used to analyze the overall structure and evolution characteristics of the innovation cooperation network (Table 1).

**Table 1.**
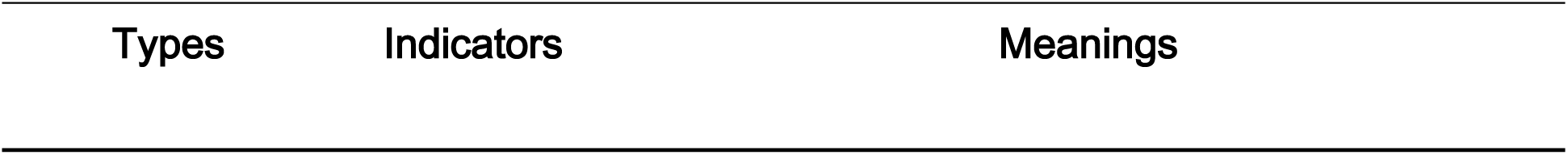
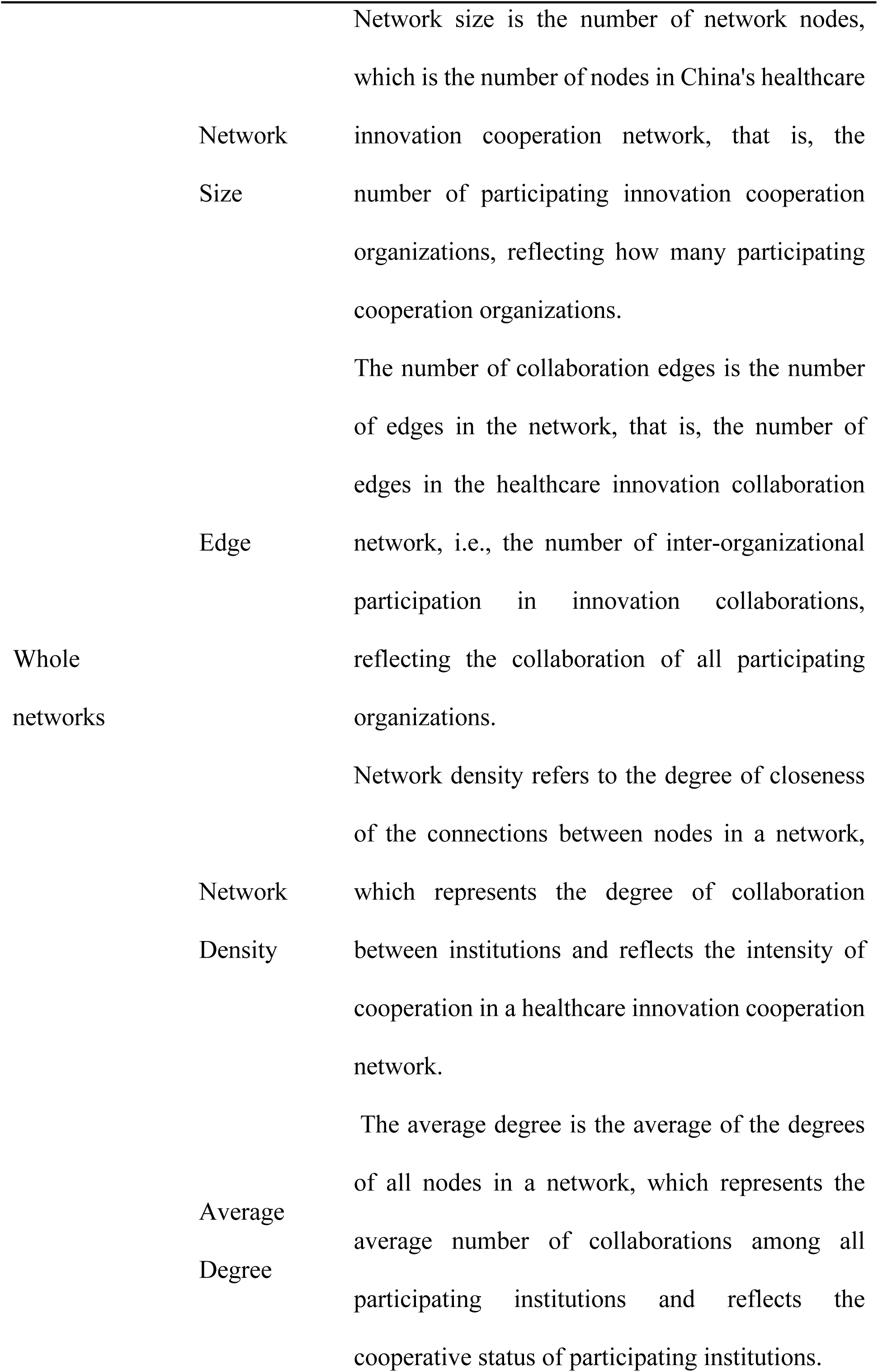
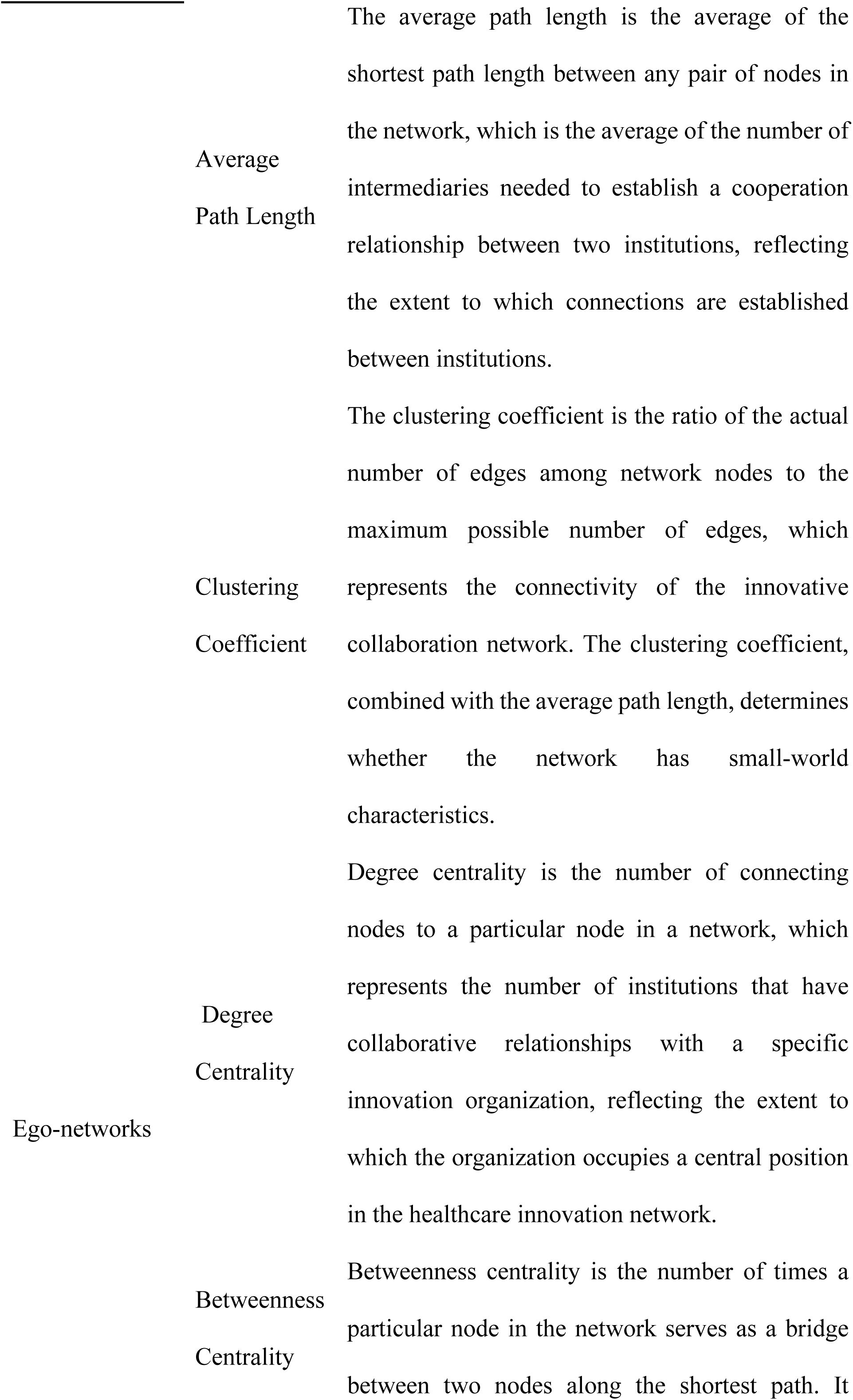
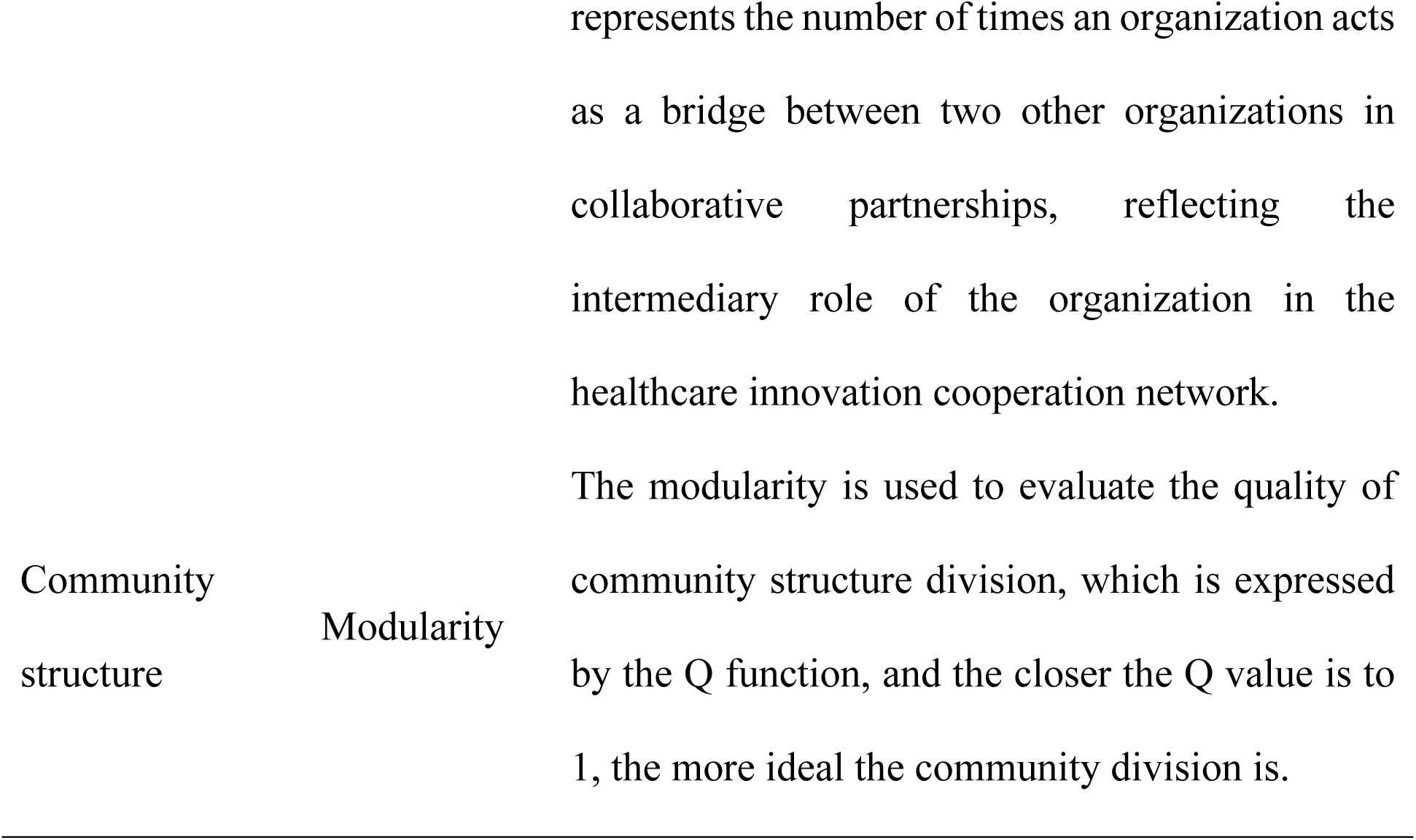
Basic indicators and meanings of China’s healthcare innovation cooperation network.

### Analysis of healthcare innovation collaboration in China

#### Trend analysis of innovation cooperation

To analyze the status of innovation cooperation in the field of medical and health care, this paper counts the percentage of projects and cooperation projects of the Chinese Medical Science and Technology Award every year (Fig.1).

**Fig. 1.**
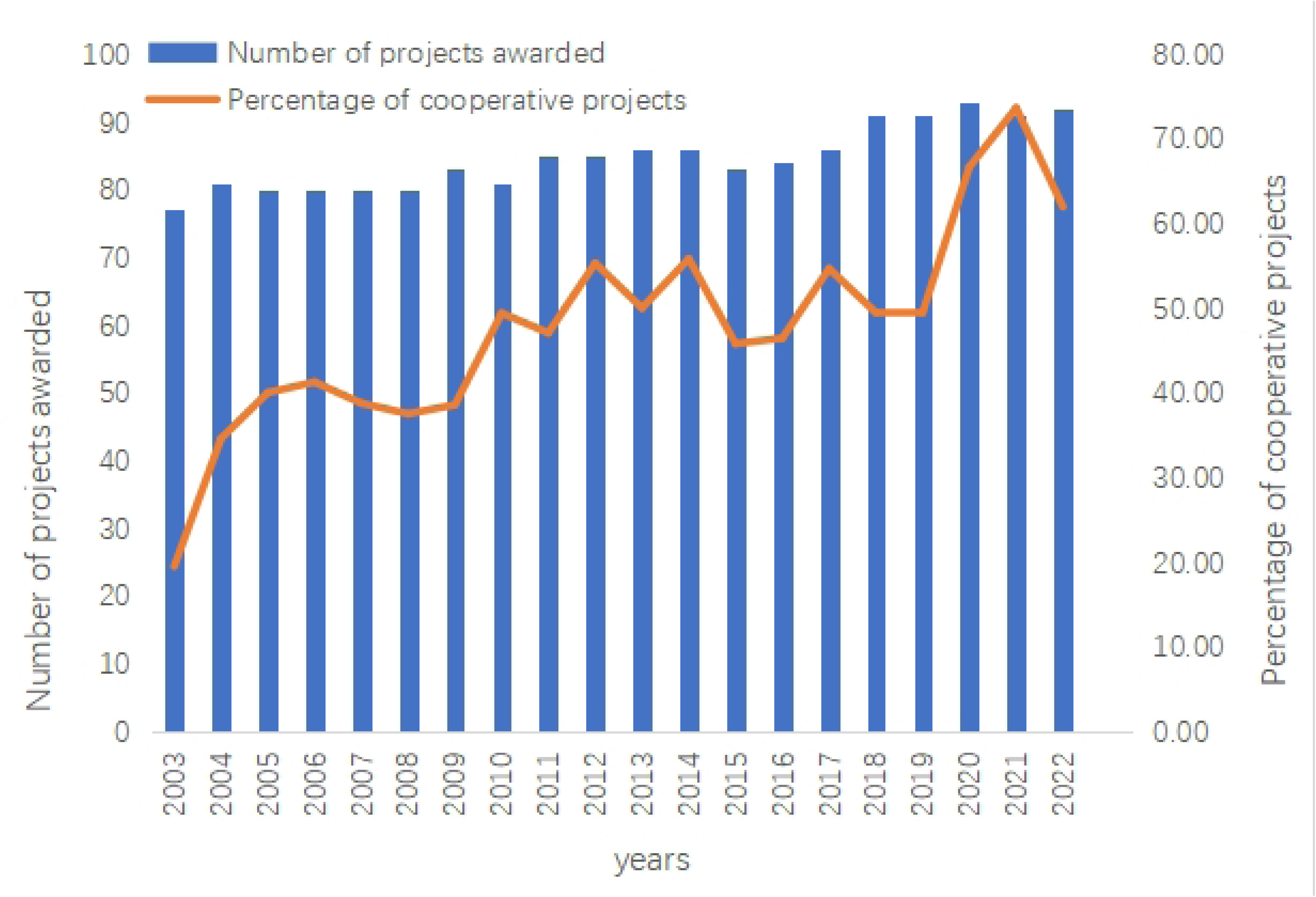
Annual distribution of the Chinese Medical Science and Technology Award projects and cooperative projects

In Fig.1, from 2003 to 2022, the number of annual awards of the Chinese Medical Science and Technology Award increased slightly, but the overall number remained stable and balanced; the proportion of cooperative projects exceeded 50% by 2010, and from 2003 to 2010, the proportion of cooperative projects showed a significant growth trend. From 2011 to 2019, the proportion of cooperative projects showed alternating growth and decrease, but the overall proportion of cooperation was around 50%. Since 2019, the proportion of cooperative projects again has increased significantly, of which the proportion is over 70% in 2021. On the whole, it shows that more and more organizations are carrying out innovative activities through cooperation, and cooperation has become an important way to promote innovation and development in the field of healthcare.

### Analysis of medical and health innovation cooperation based on social network analysis

In this paper, taking institutions as network nodes and inter-institutional cooperation as edges, Gephi is used to construct a medical and health innovation cooperation network, and the structure and evolution of China’s medical and health innovation cooperation network are analyzed from three aspects: the overall characteristics of the network, the individual characteristics of the network, and the community structure. The data is divided into four stages according to five-year intervals, which are 2003-2007, 2008-2012, 2013-2017, and 2018-2022. Meanwhile, Using Gephi software to draw the network mapping of the four stages of medical and health innovation cooperation in China (Fig.2), the nodes represent the innovation institutions, the node size represents the point degree of centrality, the larger the node, the more the institution participates in the cooperation. Besides, the network connection represents the cooperative relationship between the innovation institutions, and the thicker the edges indicate that the more the number of awards won for the cooperation between the two innovation institutions is, the closer the cooperative relationship is.

**Fig. 2.**
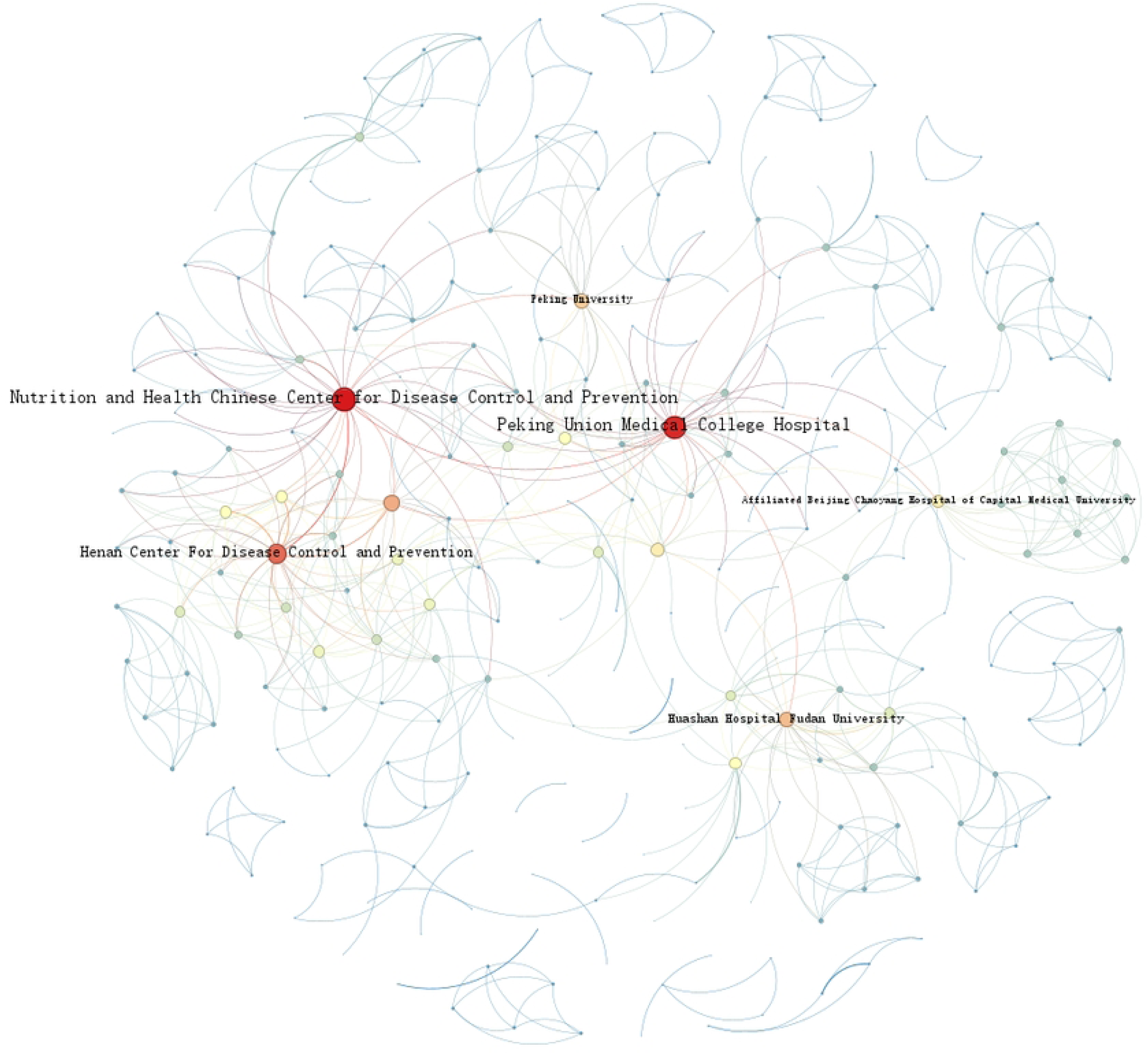
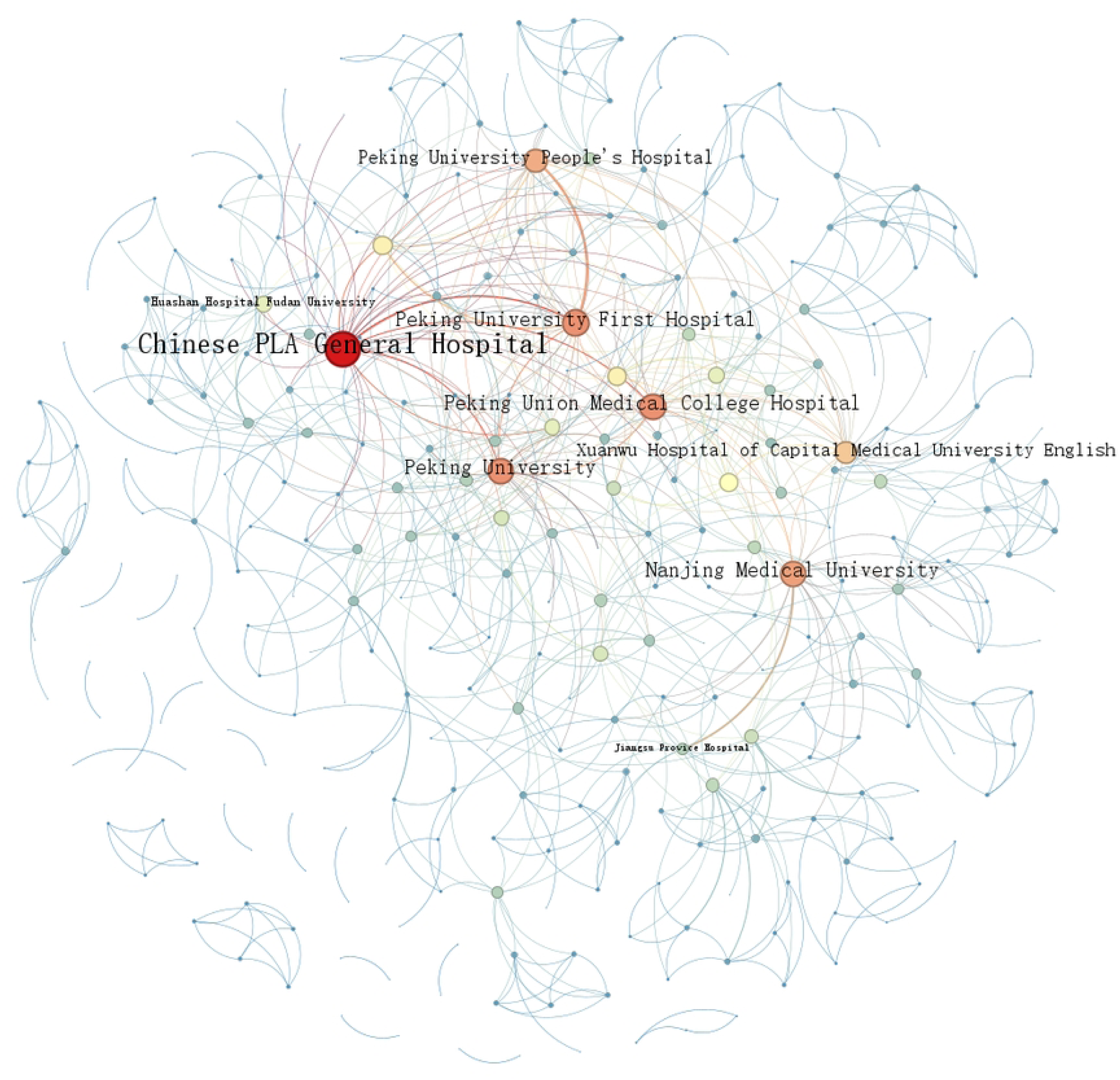
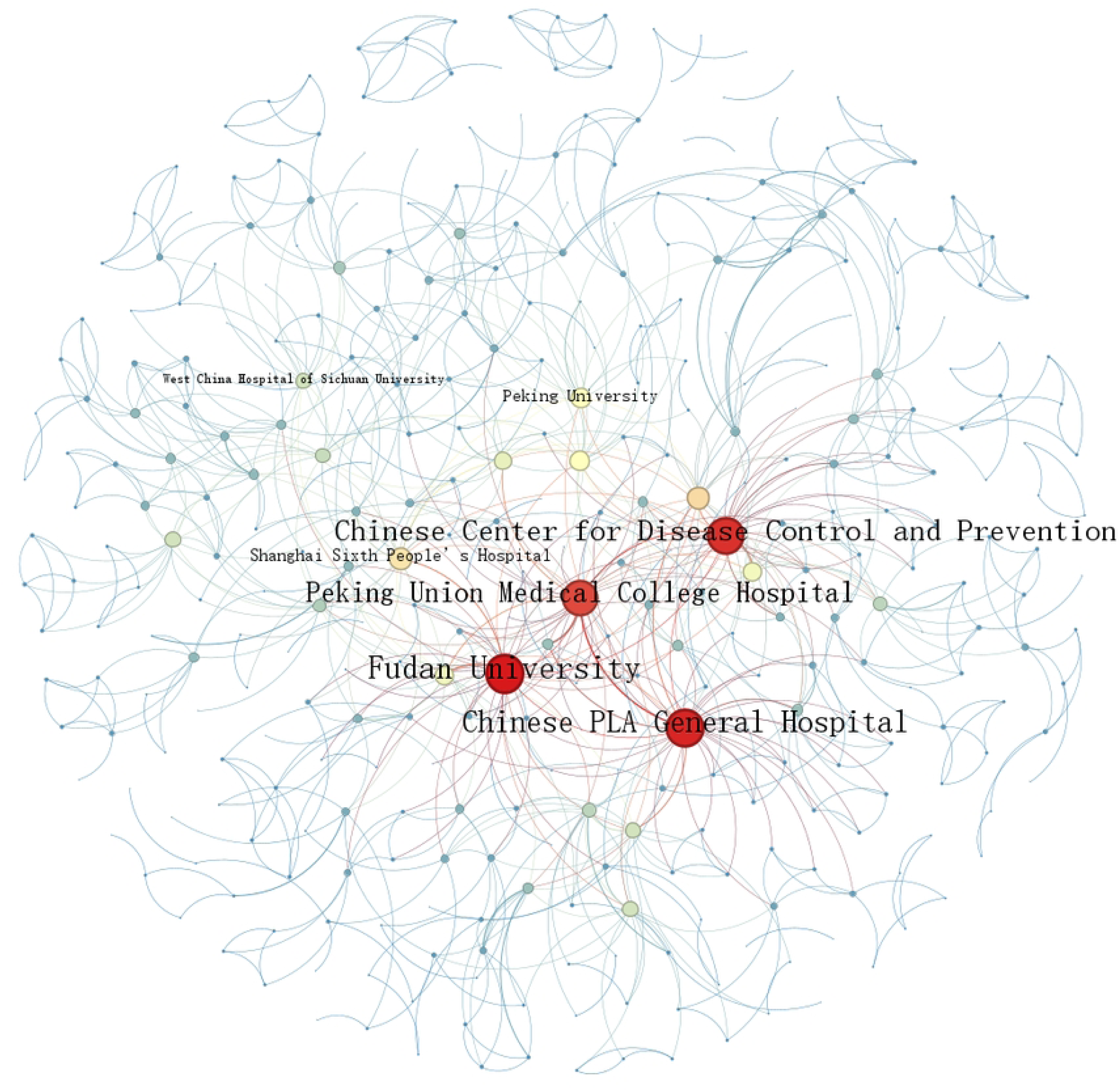
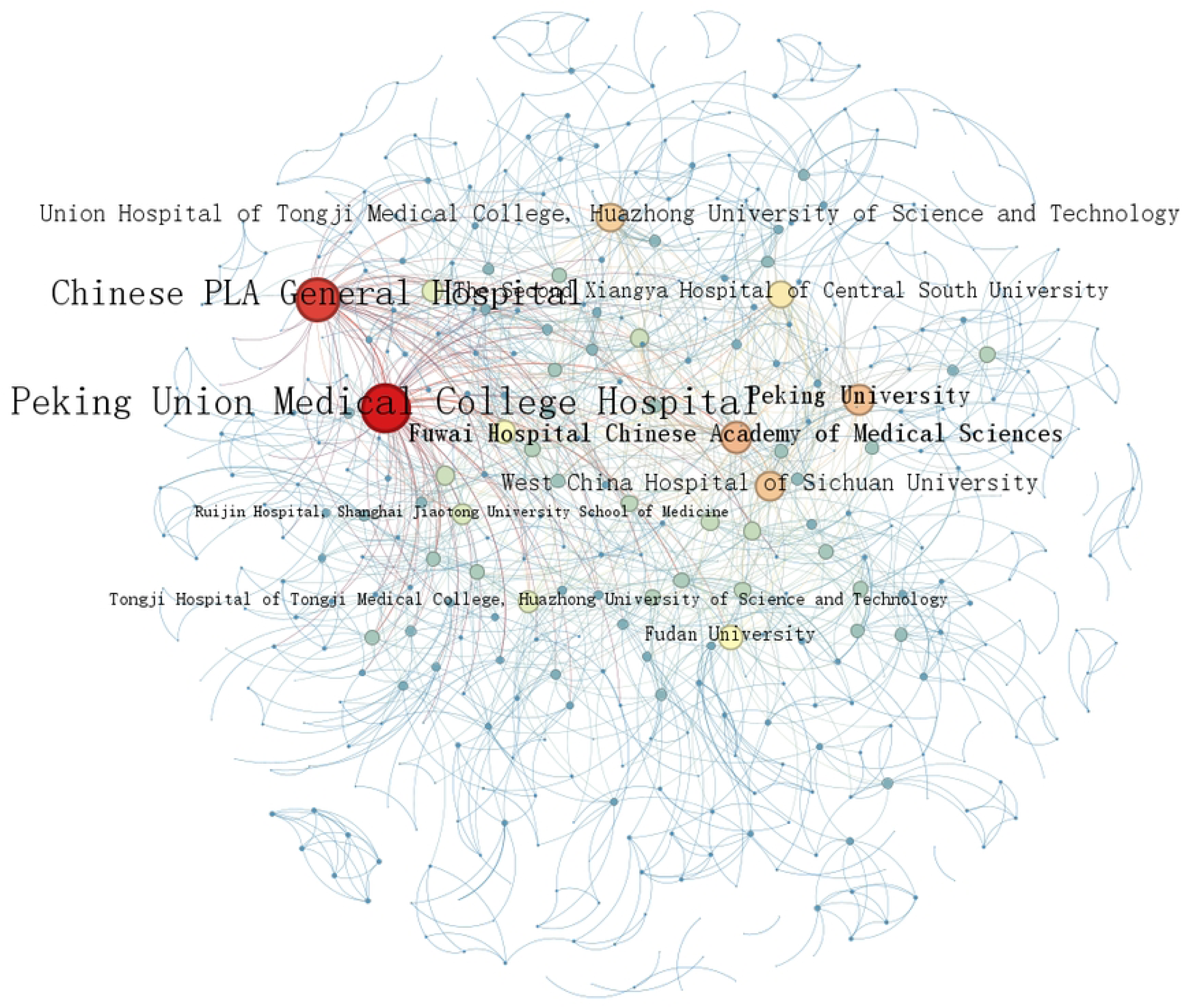
Innovation cooperation network of healthcare organizations(a)Year 2003-2007 (b) Year 2008-2012 (c) Year 2013-2017 (d) Year 2018-2022

To effectively characterize the structural features of the network at different stages, this paper uses Gephi to calculate the overall structural indicators of the healthcare innovation cooperation network at each of the four stages (Table 3).

Combined with Fig.2 and Table 2, we compare and analyze the performance of the overall basic characteristics of China’s healthcare innovation cooperation network at different stages:

(1) The scale of the innovation cooperation network continues to expand. With the continuous development of innovation activities, more and more innovation institutions are involved in innovation cooperation. In terms of the time dimension, the number of institutions involved in innovation cooperation has increased from 263 in 2003-2007 to 461 in 2018-2022, and the number of cooperation edges has increased from 642 to 1,341; in terms of the growth trend, the scale of innovation institutions increases by 20%, 16%, and 27% in 2008-2012, 2013 -2017, and 2018-2022, and the number of cooperative edges increases by 23%, 9%, and 56% respectively; the average degree also shows an overall increasing trend. The trends of network size, cooperation edges and average degree indicate increasing cooperation between institutions in the healthcare field and increasing depth of cooperation.
(2) The network structure is sparse and institutional innovation cooperation needs to be strengthened. With the continuous expansion of the scale of the innovation cooperation network, and the growth of innovation cooperation is lower than the increase in the number of innovation institutions, the network density shows a downward trend, from 0.019 in 2003-2007 to 0.013 in 2013-2017 and 2018-2022. The overall network density is low, indicating that the cooperative relationship between institutions is relatively loose and sparse, which shows that there is still much room for the development of collaborative relationships among innovation institutions.

**Table 2.** Indicators of the structure of China’s healthcare innovation cooperation network.

|  | 2003-2007 | 2008-2012 | 2013-2017 | 2018-2022 |
| --- | --- | --- | --- | --- |
| Network Size | 263 | 315 | 364 | 461 |
| Edge | 642 | 792 | 862 | 1 341 |
| Average Degree | 4.882 | 5.029 | 4.736 | 5.818 |
| Network Density | 0.019 | 0.016 | 0.013 | 0.013 |
| Average Path Length | 4.004 | 3.924 | 4.047 | 3.737 |
| Clustering Coefficient | 0.806 | 0.712 | 0.768 | 0.749 |

The characteristics of the small-world network are obvious, and the flow of innovative resources is efficient. "Smaller average path length and higher clustering coefficient" can determine the small-world characteristics of innovation cooperation networks. The clustering coefficient of the healthcare innovation cooperation network stays between 0.712 and 0.806, with a high degree of network aggregation; the average path length ranges from 4.007 to 3.737, and a link can be created between any two innovation organizations through 3-4 intermediary organizations. The above results show that the healthcare cooperation network presents obvious small-world characteristics, which are conducive to high-speed circulation and the efficient transmission of innovation resources in the cooperation network.

#### Analysis of individual network characteristics

To effectively characterize the position of institutions in the healthcare innovation cooperation network, this paper uses Gephi to calculate the point degree centrality (DC) (Table 3) and the mediator centrality (BC) (Table 4) of the top ten ranked institutions in the four stages.

**Table 3.** Key nodes of China’s healthcare innovation network and their point degree centrality.

| Table 3 Key nodes of China's healthcare innovation network and their point degree centrality (DC) |  |  |  |  |
| --- | --- | --- | --- | --- |
| DC | 2013-2017 | DC | 2018-2022 | DC |
| 35 | Fudan University | 39 | Peking Union Medical College Hospital | 56 |
| 26 | Chinese PLA General Hospital | 38 | Chinese PLA General Hospital | 51 |
| 26 | Chinese Center for Disease Control and Prevention | 37 | Fuwai Hospital Chinese Academy of Medical Sciences | 37 |
| 26 | Peking Union Medical College Hospital | 35 | Peking University | 36 |
| 25 | Peking University People's Hospital | 23 | West China Hospital of Sichuan University | 35 |
| 24 | Shanghai Sixth People's Hospital | 22 | Union Hospital of Tongji Medical College, Huazhong University of Science and Technology | 34 |
| 22 | Peking University | 20 | The Second Xiangya Hospital of Central South University | 31 |
| 19 | Affiliated Beijing Chaoyang Hospital of Capital Medical University | 20 | Fudan University | 29 |
| 19 | Shanghai Jiao Tong University | 19 | Peking University People's Hospital | 28 |
| 18 | National Institute for Viral Disease Control and Prevention Chinese Center for Disease Control and Prevention | 19 | Tsinghua University | 25 |

**Table 4.**
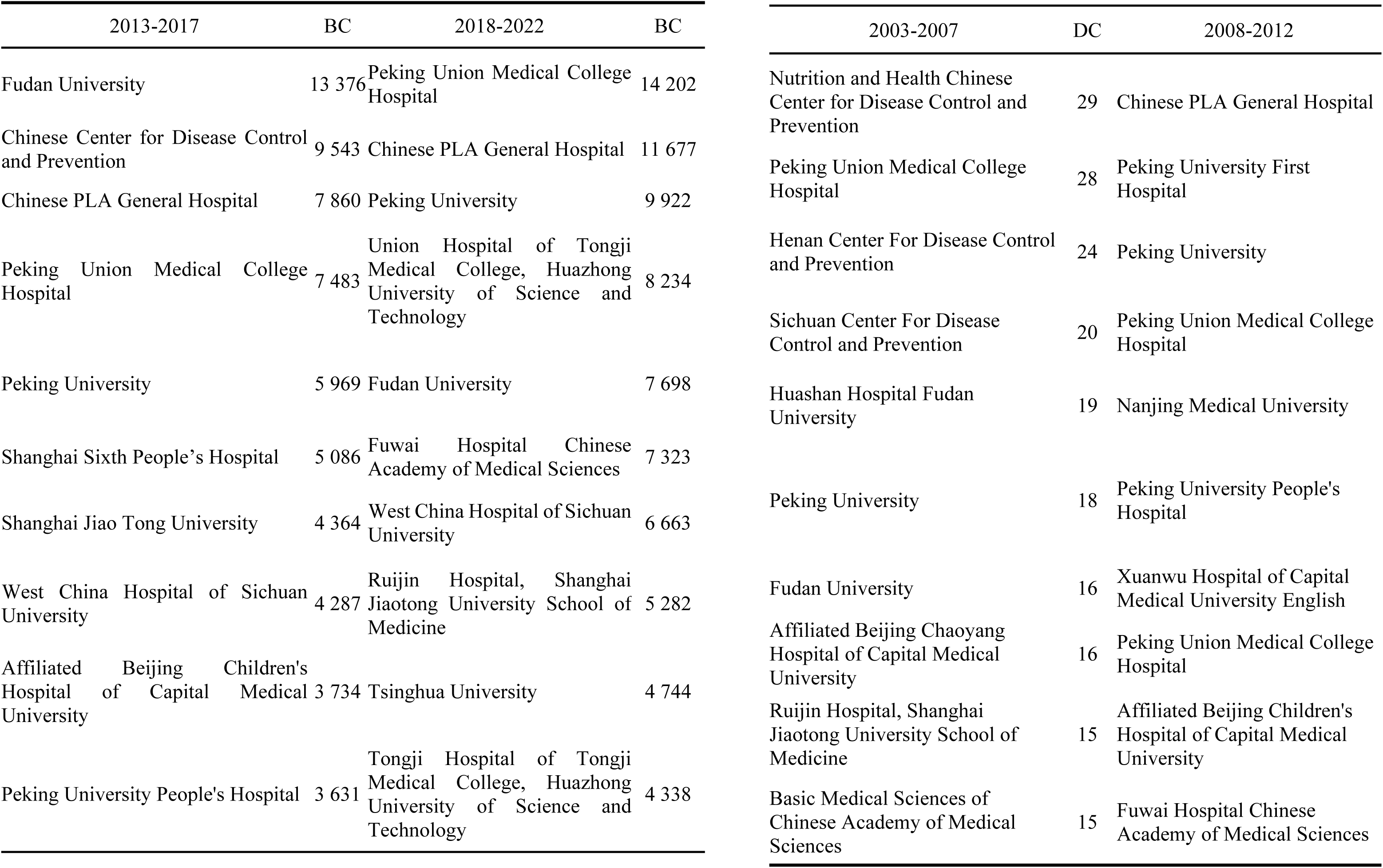
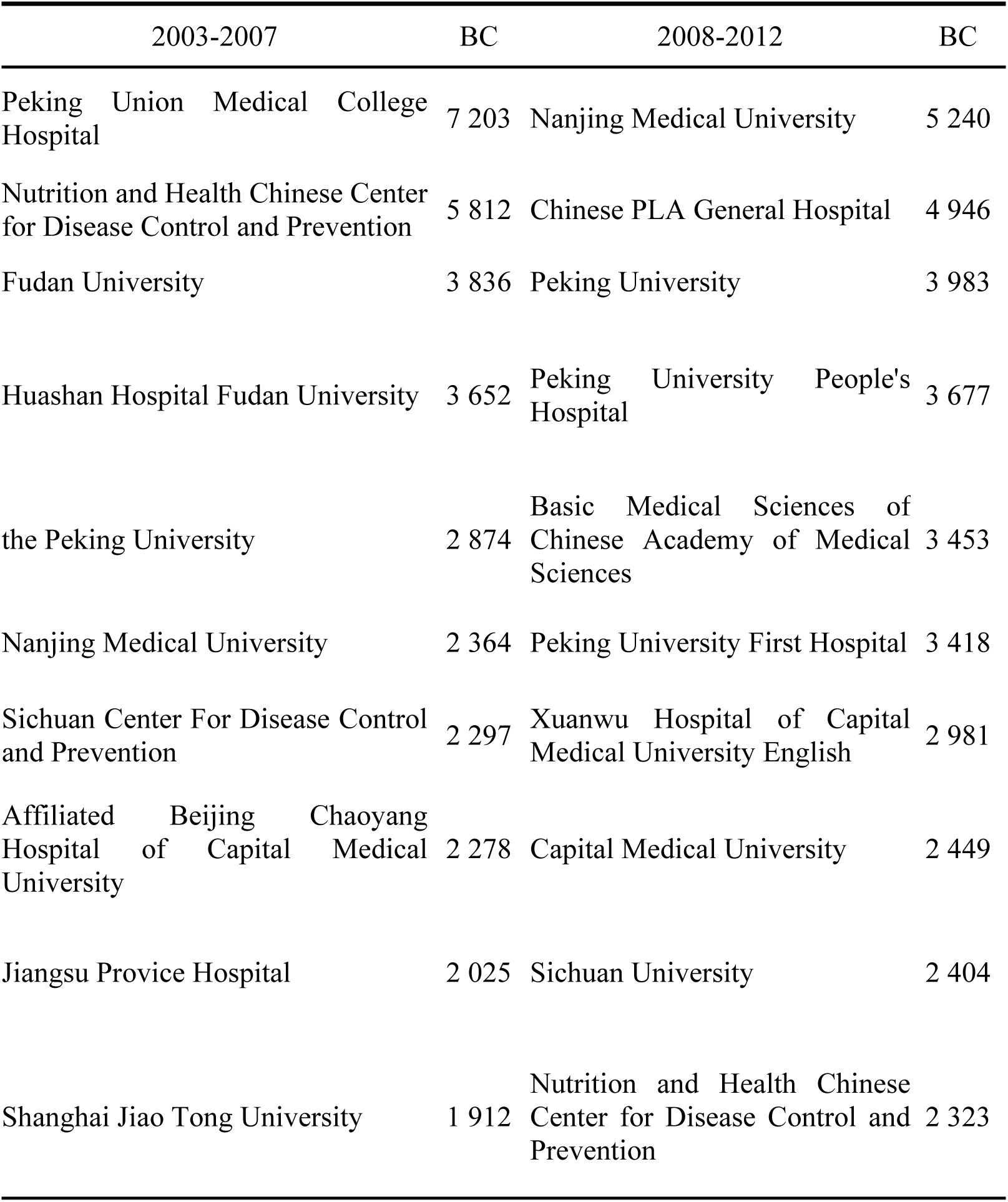
Key nodes of China’s healthcare innovation network and their betweenness.

In Tables 3 and 4, the performance of degree centrality and betweenness centrality of the main institutions of our healthcare innovation cooperation network from 2003 to 2022:

(1) From the time dimension, the degree centrality and betweenness centrality of the partner institutions showed a significant growth trend. The degree centrality of the Institute of Nutrition and Health Chinese Center for Disease Control and Prevention (CDC) was the highest at 29 in 2003-2007, and the degree centrality of Fudan University, which ranked eighth in 2018-2022, was also 29; The Peking Union Medical College Hospital had the highest betweenness centrality of 7,2023 in 2003-2007, and the sixth-ranked Fuwai Hospital Chinese Academy of Medical Sciences had an betweenness centrality of 7,323 in 2018-2022.The degree centrality and betweenness centrality indicate that the innovation institutions have strengthened their cooperation connection, that the core of the cooperative institutions in the innovation cooperation network is increasing, and that the ability to control the flow and transmission of innovation resources in the network is increasing.
(2) In terms of the type of core structure, regardless of degree centrality or intermediary centrality, hospitals are in the core position in the innovation network, and the Chinese PLA General Hospital, the Peking Union Medical College Hospital, and the Peking University People’s Hospital have a higher degree centrality and intermediary centrality, which indicates that hospitals are actively involved in inter-institution innovation cooperation, and have a stronger ability to regulate resources in the whole innovation cooperation network. Colleges and universities are more prominent in the betweenness centrality degree, and Peking University and Fudan University rank in the top ten in all stages, indicating that colleges and universities play a key role in bridge linkage in the whole innovation cooperation network. In the first three stages, government-funded research institutes are in a key position in the innovation cooperation network, because the Chinese Center for Disease Control and Prevention and the Chinese Academy of Medical Sciences are involved in the research of medical and health innovation cooperation as cooperating institutes, indicating that the government also plays a key role in the healthcare innovation network collaboration network. None of the pharmaceutical companies appear in the top ten rankings in terms of point degree centrality or intermediary centrality, indicating that the companies are in a peripheral position in the innovation collaboration network.

Overall, the control ability of the core nodes in China’s medical and health innovation cooperation network continues to increase, hospitals are in an absolute central position in the innovation cooperation network, universities play a prominent role in innovation cooperation, the role of government-funded research institutions has a downward trend, and enterprises are at the edge of the innovation cooperation network.

#### Analysis of community structure

Community structure is a collection of closely connected innovation subjects in an innovation network, and the evolutionary characteristics are of great significance for understanding the organizational structure and revealing the potential relationships of the network [15]. To analyze the community structure and evolution of China’s medical and health innovation cooperation network, this paper uses Gephi’s modular processing to generate the community structure (Fig.3).

**Fig. 3.**
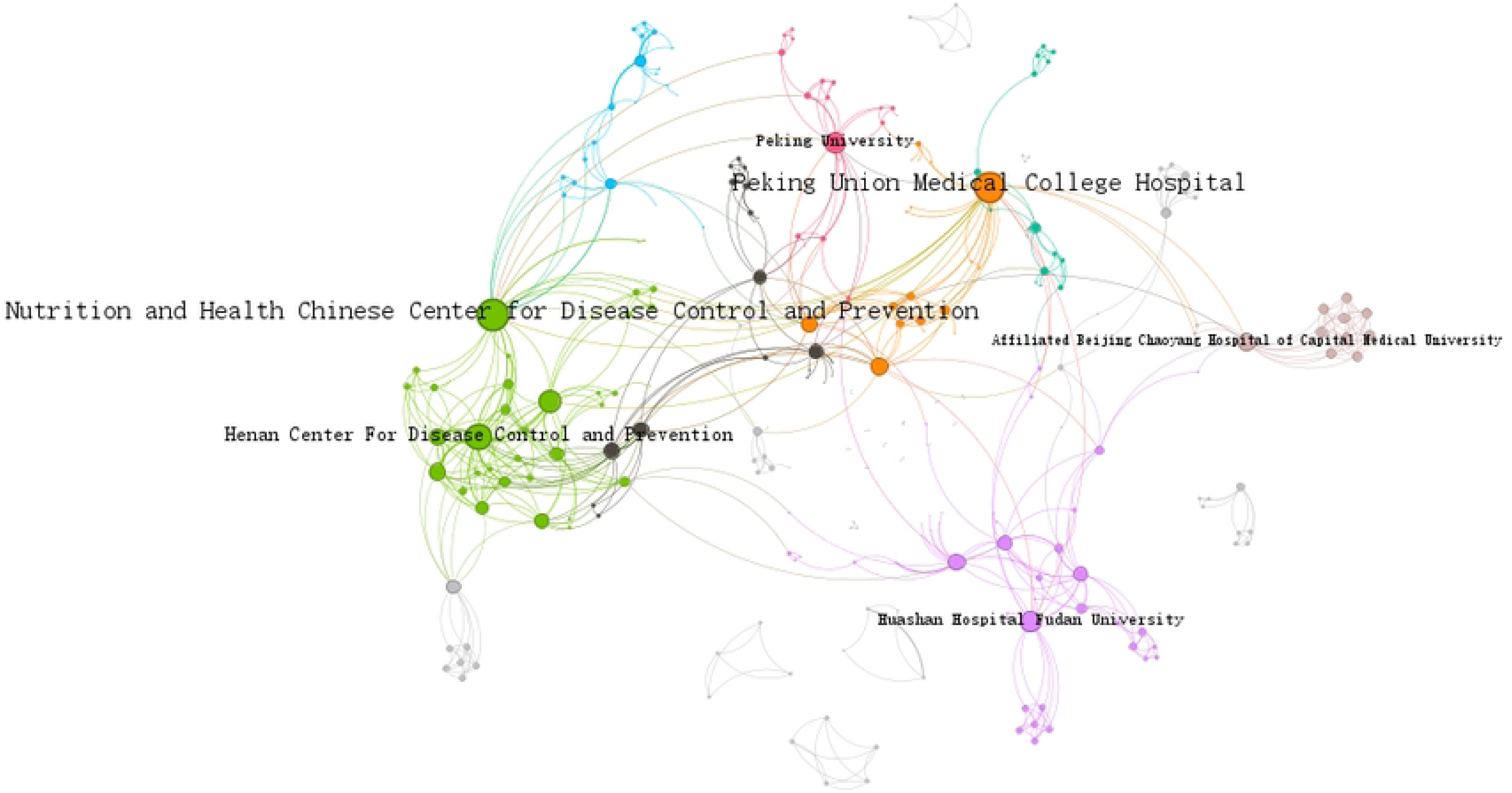
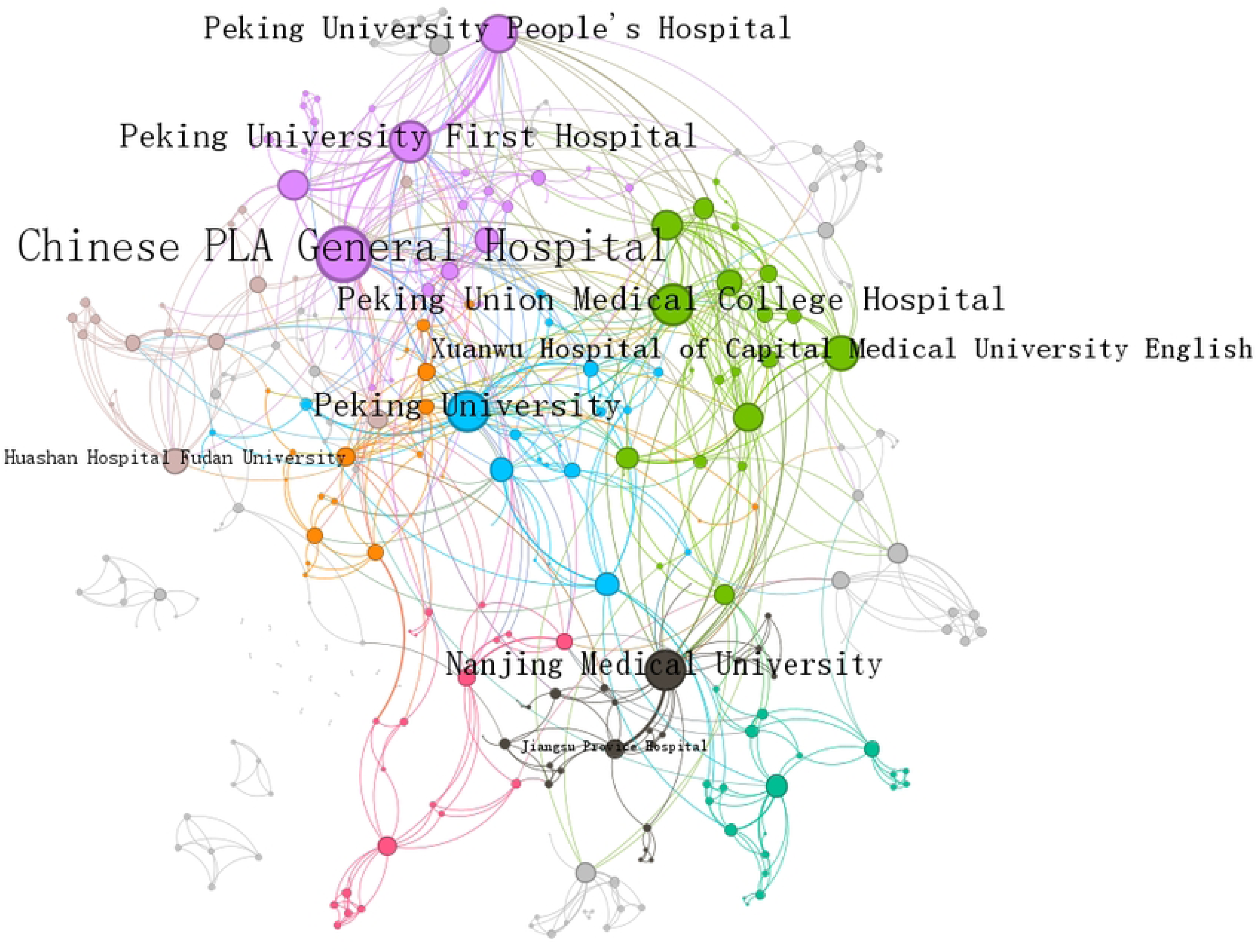
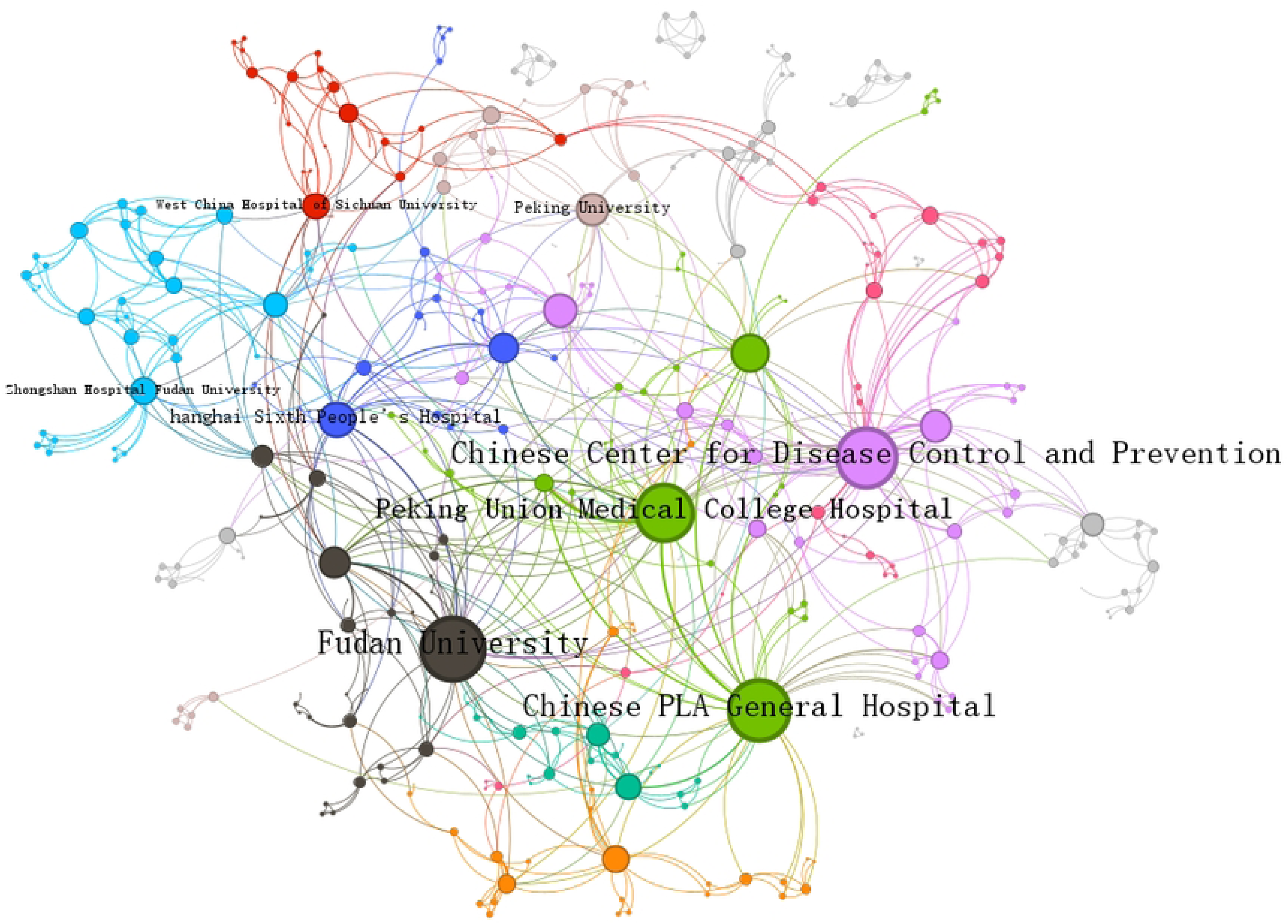
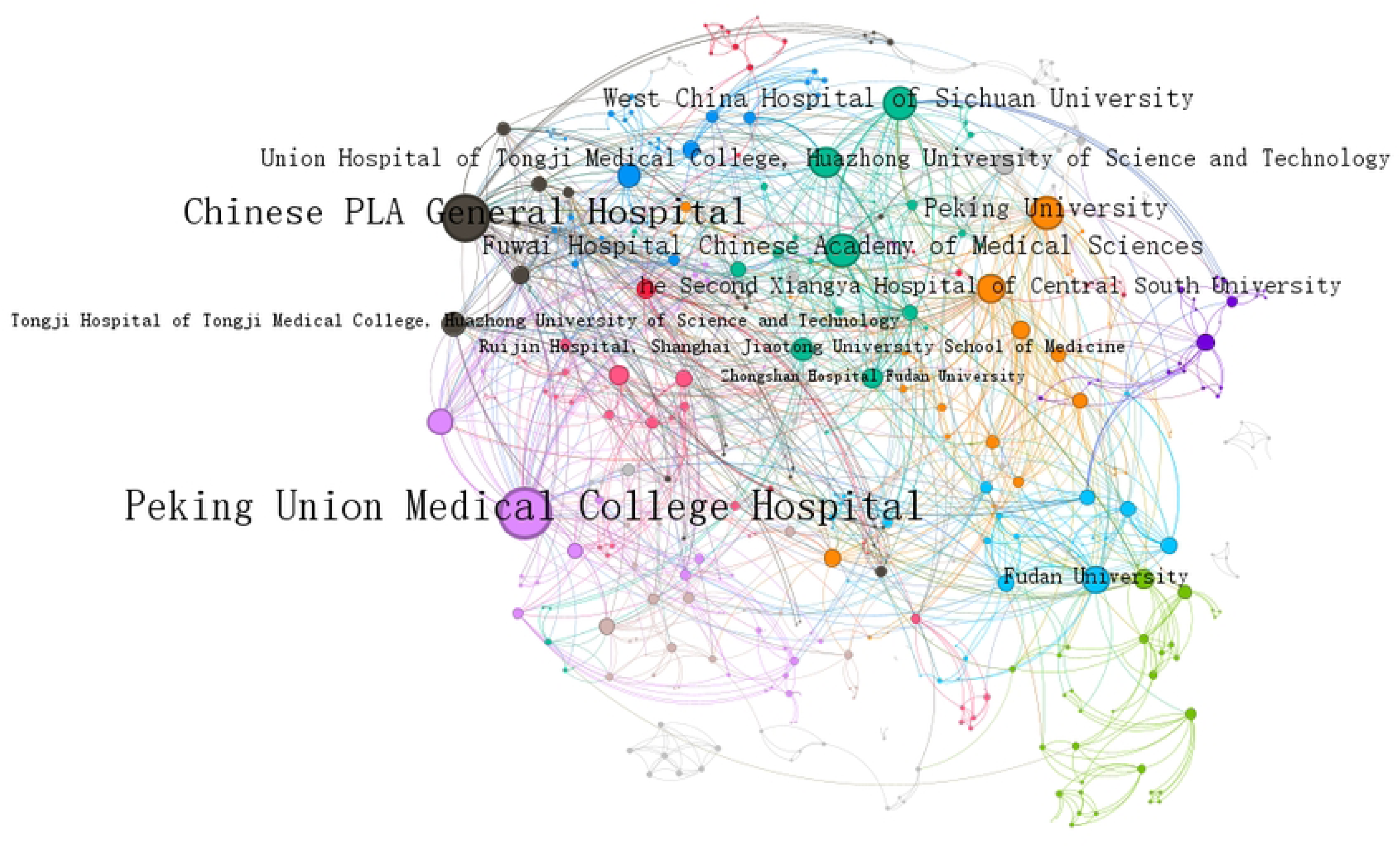
The community structure of China’s medical and health innovation cooperation network(a)Year 2003-2007 (b) Year 2008-2012 (c) Year 2013-2017 (d) Year 2018-2022

In Fig.3, the nodes of the same color in each stage of the healthcare innovation cooperation network indicate the same community structure. Overall, it indicates that the community structure of China’s healthcare innovation cooperation network in the period of 2003-2022 has become closer and closer, and the breadth and depth of cooperation have significantly increased. To analyze the community structure of the innovation cooperation network more clearly, the relevant characteristic indexes of the community structure are calculated (Table 6).

In Fig.3 and Table 5, the community structure of China’s healthcare innovation cooperation network and its evolutionary performance:

**Table 5.** China’s medical and health innovation cooperation network associations.

|  | 2003-2007 | 2008-2012 | 2013-2017 | 2018-2022 |
| --- | --- | --- | --- | --- |
| Number of associations | 28 | 32 | 29 | 27 |
| Maximum number of association nodes | 40 | 38 | 42 | 54 |
| Minimum number of association nodes | 2 | 2 | 2 | 2 |
| Modularity | 0.766 | 0.673 | 0.696 | 0.612 |

(1) Significant community structure

As shown in Fig.3, the network scale and number of cooperation edges are the same as in Fig.2, and the number of community structures in each stage remains stable and balanced. The number of nodes of the largest community is stable in the first three stages, and there is a large increase in the fourth stage; the modularity decreases from 0.766 to 0.612, and the overall level remains above 0.6. The above shows that the quality of community division is good, the spatial organization structure of associations is obvious, the internal community cooperation is closer, and there is also cooperation and exchange between communities.

(2) Visible nodes of larger association centers

In Fig.3, there are larger nodes within the larger community, indicating the existence of one or more community centers. From 2003 to 2012, most of the inter-institutional cooperation was mainly based on the formation of communities formed in close geographical proximity, such as the cooperative community centered on Peking University and the Peking University People’s Hospital, and the cooperative community formed by Fudan University and hospitals affiliated to Shanghai Jiao Tong University School of Medicine etc.; indicating that there is a larger resistance to inter-institutional cooperation in the exchange of knowledge and the sharing of resources in this stage, resulting in a limited ability of institutional innovation cooperation to radiate. From 2013 to 2022, the award-winning institutions are no longer confined to the same region, forming innovative cooperative community centered on West China Hospital of Sichuan University, Union Hospital of Tongji Medical College, Huazhong University of Science and Technology, and Xiangya Hospital of Central South University, Sun Yat-sen University, and the First Affiliated Hospital of Sun Yat-sen University, which have gradually grown up to become the core of the communities, indicating that the depth of inter-institutional innovative cooperation has been increased and the radiation scope of innovation cooperation network has been more extensive.

The possible reasons for the formation of such communities are that hospitals have rich clinical research resources, which are an important source for the development of healthcare innovation, and colleges and universities have more innovation resources, which provide a constant flow of talent for healthcare innovation. Hospitals and universities are more responsible for information dissemination and results diffusion in China’s healthcare innovation cooperation network, and have greater control in the network, further proving the importance of hospitals and universities in the innovation cooperation network.

#### Analysis of regional cooperation

To analyze the status of regional cooperation in cooperation, this paper provides statistics on intra-regional and inter-regional cooperation in China’s provinces (municipalities directly under the central government, autonomous regions, and special administrative regions) and other countries and regions (Table 2).

As can be seen from Fig.4, China’s regional innovation cooperation in the field of medical and health care not only widely covers domestic provinces, cities and autonomous regions, but also cooperates with the United States, South Korea, Japan, Australia and other countries; both intra-regional and inter-regional cooperation is relatively strong in the eastern region, while the cooperation in the central region is dominated by inter-regional cooperation, and both intra-regional and inter-regional cooperation in the western region is relatively weak.

**Fig. 4.**
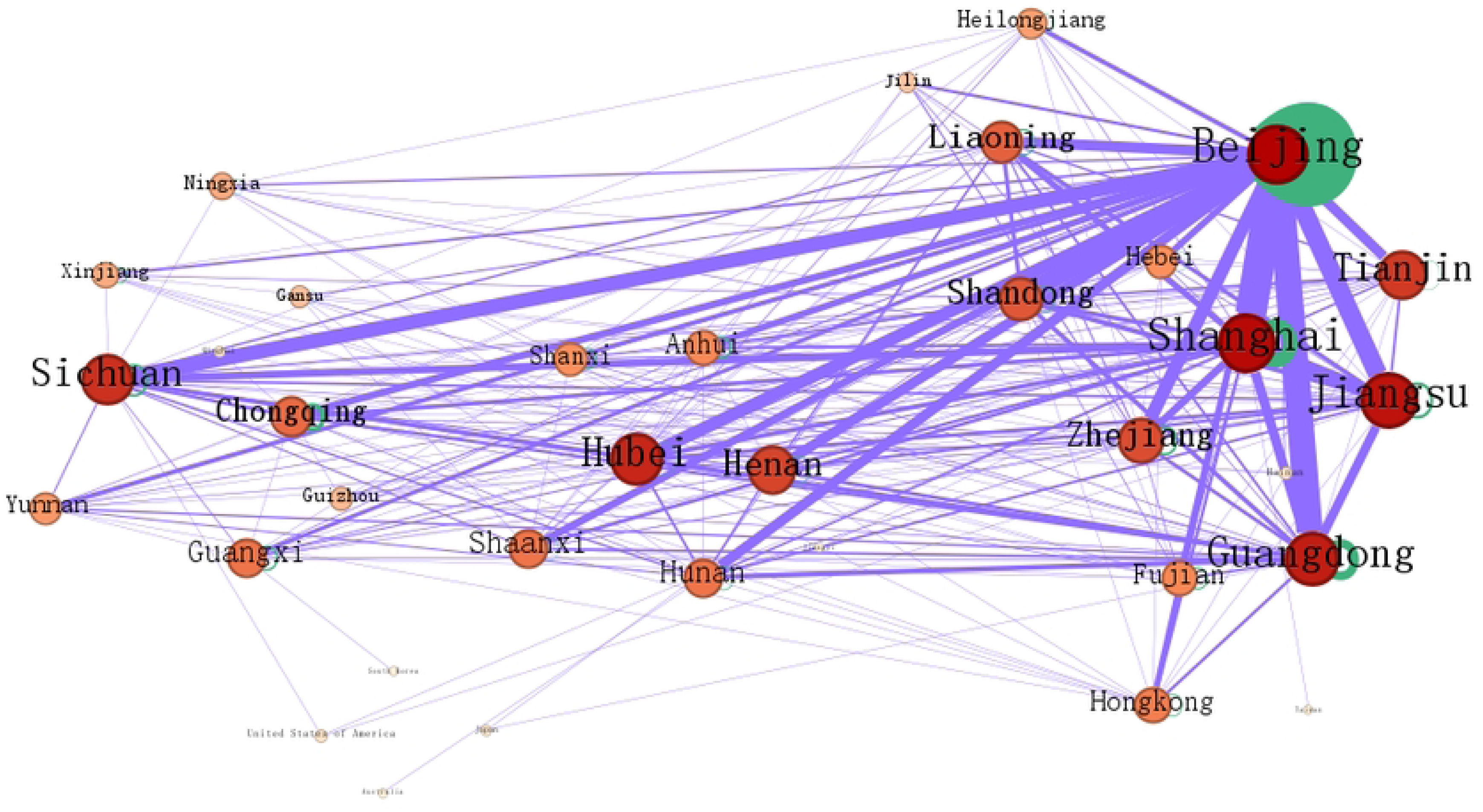
The regional innovation cooperation network of China’s medical and health innovation cooperation network

### Conclusions and recommendations

#### Conclusions

Taking the institutional cooperation of the award-winning projects of Chinese Medical Science and Technology Award from 2003 to 2022 as the research object, this paper firstly analyzes the basic situation of institutional innovation cooperation in the field of healthcare; then constructs the institutional innovation cooperation network; and finally explores the structural law and evolutionary characteristics of China’s healthcare innovation cooperation network, and draws the following conclusions: (1) Basic characteristics of innovation cooperation. The participation of institutions in innovation cooperation has shown a rapid growth trend and gradually become an important way of innovation. (2) Overall characteristics of innovation cooperation network. The scale of the innovation cooperation network and the number of cooperation edges continue to expand and grow; the small-world effect of the innovation cooperation network is more obvious, which is conducive to the circulation and exchange of innovation resources; however, the whole innovation cooperation network has a low network density and sparse network structure, indicating that there is still considerable room for institutional innovation cooperation. (3) Individual characteristics of innovation cooperation network. From the time dimension, the degree centrality and betweenness centrality of the core nodes in the innovation cooperation network show a larger growth trend, indicating that the position in the network is becoming more and more important; from the type of institutions, hospitals are in the absolute core position in the innovation network by their rich clinical research resources; colleges and universities have rich human resources and play a bridging role in the innovation cooperation network; government-funded research institutions and government-funded research institutions and enterprises are less likely to appear in the core of the innovation network and more often at the edge of the network. (4) Characteristics of innovation cooperation network communities. The innovation cooperation network has obvious community characteristics; larger communities have obvious community centers, and most of them are dominated by hospitals, colleges and universities, reflecting their leading and radiating role in the innovation cooperation network. (5) The development of regional innovation cooperation is seriously unbalanced, with a higher degree of institutional participation in cooperation in the eastern region and a lower degree of participation in cooperation in the central and western regions

#### Policy recommendations

The analysis of China’s healthcare innovation cooperation shows that cooperation has become an important way of innovation, but there are still problems such as unbalanced, low network density and insufficient cooperation in the healthcare innovation cooperation network. To comprehensively promote China’s inter-institutional innovation cooperation in the field of healthcare, the following policy recommendations are proposed:

(1) Strengthen the circulation and exchange of innovation resources among regions, and actively build an innovation cooperation system for balanced regional development. Regions with strong innovation cooperation capacity, such as Beijing, Shanghai, Guangdong and Jiangsu, should not only deeply explore inter-institutional cooperation within the region, but also actively unite with central and western regions to carry out inter-regional innovation cooperation, thus enhancing the capacity of central and western regions to participate in innovation cooperation; central and western regions need to integrate local advantages into the innovation cooperation network according to their actual conditions, improve their innovation cooperation capacity, and enhance intra-regional cooperation on this basis. Reasonable allocation of innovation resources, encouragement of innovation exchange and cooperation among regions, narrowing the gap of regional innovation cooperation, and optimizing the balanced development of the innovation cooperation network in the field of medical and healthcare towards the region.
(2) Actively give full play to the radiation-driven role of core institutions in the innovation cooperation network, and build a robust innovation cooperation network ecology. There are institutions at the core position in both the entire innovation cooperation network and larger communities, such as the Chinese PLA General Hospital, Peking University and its affiliated hospitals, Fudan University, etc. Actively exerting the radiation-driven ability of these institutions in innovation cooperation, promoting the efficient circulation of innovation resources in the cooperation network, and realizing the transformation of the cooperation network from "big" to "strong".
(3) Actively promote the role of enterprises in innovation cooperation networks and build a robust innovation cooperation network structure supported by a diversified core. Given the relatively weak position of enterprises in the whole innovation cooperation network, it is necessary to increase the motivation of enterprises to participate in innovation cooperation from the aspect of the policy, promote diversified innovation cooperation modes, cultivate a group of enterprises that can be at the core of the innovation network, and deepen the integration of innovation cooperation among multiple subjects, to enhance the network density of the innovation cooperation network, and build a stable innovation network structure.

## Data Availability

The data underlying the results presented in the study are available from Chinese Medical Association (https://www.cma.org.cn/col/col38/index.html ).

## Acknowledgments

We would like to express our deepest gratitude to the organizations that provided the relevant data files.

